# Appropriate management of steroids and discharge planning during and after hospital admission for moderate-severe ulcerative colitis

**DOI:** 10.1101/2022.01.24.22269684

**Authors:** Parambir S. Dulai, Victoria Rai, Laura E. Raffals, Dana Lukin, David Hudesman, Gursimran S. Kochhar, Oriana M. Damas, Jenny S. Sauk, Alexander N. Levy, M. Anthony Sofia, Anne Tuskey, Parakkal Deepak, Andres J. Yarur, Anita Afzali, Ashwin N Ananthakrishnan, Raymond K. Cross, Stephen B. Hanauer, Corey A. Siegel

**Affiliations:** Division of Gastroenterology and Hepatology, Northwestern University, Chicago, IL; Department of Cellular and Molecular Physiology, Yale University, New Haven, CT; Division of Gastroenterology and Hepatology, Mayo Clinic, Rochester, MN; Division of Gastroenterology and Hepatology, Weill Cornell Medicine, New York, NY; Division of Gastroenterology and Hepatology, New York University, New York, NY; Division of Gastroenterology and Hepatology, Alleghany Health, Pittsburgh, PA; Division of Gastroenterology and Hepatology, University of Miami Miller School of Medicine, Miami, FL; Division of Gastroenterology and Hepatology, University of California Los Angeles, Los Angeles, CA; Division of Gastroenterology and Hepatology, Tufts Medical Center, Boston, MA; Division of Gastroenterology and Hepatology, Oregon Health & Science University, Portland, OR; Division of Gastroenterology and Hepatology, University of Virginia, Charlottesville, VA; Division of Gastroenterology and Hepatology, Washington University School of Medicine, St. Louis, MO; Division of Gastroenterology and Hepatology, Wisconsin University, Milwaukee, WI; Division of Gastroenterology and Hepatology, Ohio State University, Columbus, OH; Division of Gastroenterology and Hepatology, Massachusetts General Hospital, Boston, MA; Division of Gastroenterology and Hepatology, University of Maryland School of Medicine, Baltimore, MD; Section of Gastroenterology and Hepatology, Dartmouth-Hitchcock Medical Center, Lebanon, NH

**Keywords:** ulcerative colitis, steroids, hospital, discharge

## Abstract

**Background:** Limited guidance exists for the post-discharge care of ulcerative colitis (UC) patients hospitalized for moderate-severe flares.

**Methods:** RAND methodology was used to establish appropriateness of inpatient and post-discharge steroid dosing, discharge criteria, follow-up, and post-discharge biologic or small molecule initiation. A literature review informed the panels voting, which occurred anonymously during two rounds before and after a moderated virtual session.

**Results:** Methylprednisolone 40-60mg IV every 24 hours or hydrocortisone 300mg IV three times daily are appropriate for inpatient management, with methylprednisolone 40mg being appropriate if intolerant of higher doses. It is appropriate to discharge patients once rectal bleeding has resolved (Mayo sub score 0-1) and/or stool frequency has returned to baseline frequency and form (Mayo sub score 0-1). It is appropriate to discharge patients on 40mg of prednisone after observing patients for 24 hours in-hospital to ensure stability prior to discharge. For patients being discharged on steroids without in-hospital biologic or small molecule therapy initiation, it is appropriate to start anti-TNF therapy after discharge for anti-TNF naïve patients. For anti-TNF exposed patients it is appropriate to start vedolizumab or ustekinumab for all patients, and tofacitinib for those with a low risk of adverse events. It is appropriate to follow up patients clinically within 2 weeks, and with lower endoscopy within 4-6 months after discharge.

**Conclusion:** We provide guidance on the inpatient and post-discharge management of UC patients hospitalized for moderate-severe flares.

**STUDY HIGHLIGHTS:** *WHAT IS KNOWN:*

- Ulcerative colitis patients hospitalized for disease flares are a high-risk population
- Guidance on evaluation and initial management during flares is provided, however, limited guidance exists on standardization of steroid management and post-discharge care

*WHAT IS NEW HERE:*

- Through a RAND Appropriateness Panel we provide guidance on the inpatient and post-discharge management of steroids, discharge criteria, post-discharge monitoring and management of biologics or small molecule therapies
- These recommendations will help to bring uniformity to care for this high-risk population, and optimize outcomes in clinical practice

## BACKGROUND

Nearly 50% of ulcerative colitis (UC) patients will require hospitalization for an acute flare at some point in their disease course.^1^ Population-based cohort estimates from the United States specifically estimate that 29% will require hospitalization within the first 5 years of diagnosis, and this increases to 39% and 49% at 10 and 20 years after diagnosis.^2^ The United States has reported a significant increase in UC-related hospitalizations between 2006 (29,888 hospitalizations; 95% CI 27,553-32,222) to 2016 (35,735 hospitalizations; 95% CI 34,284-37,186) representing a 20% increase over 10 years, and UC-related hospitalizations resulted in approximately 2 billion dollars of direct healthcare expenditures in 2016 alone.^3^ This rise in hospitalizations for UC patients in the United States has disproportionately affected minorities, those with government insurances, and those in rural centers, where care coordination may be sub-optimal due to limited resources.^3^ The need for hospitalization also represents a major event in a UC patient’s disease course, and hospitalized patients remain at a significantly increased risk for re-hospitalization, colectomy, and morbidity and mortality for up to 5 years following discharge.^1,4-8^

Despite the high-risk nature of this population, limited guidance is available for their optimal management. Steroids remain a cornerstone of treatment, however, the American College of Gastroenterology (ACG) and American Gastroenterology Association (AGA) provide differing recommendations on dosing for in-patient use of steroids, and no guidance is provided on transitioning to oral prednisone and outpatient steroid management.^9,10^ Furthermore, no societal guidelines, international organizations, or consensus statements provide guidance on discharge criteria, follow-up assessments for clinical and endoscopic activity, or post-discharge initiation of biologics or small molecules.^9-12^ Standardization of steroid management and post-discharge care for hospitalized moderate-severe UC patients is needed both for clinical practice and future clinical trials in this understudied population. To address this uncertainty, we applied the RAND Appropriateness Method to assess key scenarios most likely to influence post-discharge outcomes for UC patients hospitalized for acute moderate-severe flares.

## METHODS

### Design

The RAND Appropriateness Method uses a modified Delphi panel of specialists in combination with the best available evidence to provide appropriateness ratings for clinical scenarios.^13^ The methodology has been widely used in the field of inflammatory bowel disease (IBD) to address uncertainty around concomitant immunomodulator use with anti-tumor necrosis factor (TNF) agents,^14^ therapeutic drug monitoring,^15^ and the management of patients during the coronavirus pandemic.^16^ This iterative structured process includes a literature review presented to a specialist panel followed by 2 rounds of anonymous panel ratings, with virtual or in-person moderated discussions among participants prior to the second round of voting.

### Literature Review

A targeted literature search was performed in relation to the clinical scenarios and topic of post-hospitalization care for UC patients presenting for acute flares. Specific targeted topics included: steroid dosing in-hospital and post-discharge, in-hospital response and discharge criteria, post-discharge biologic and small molecule therapy use, and post-discharge follow-up and disease activity assessment. Guidelines and consensus recommendations from the AGA, ACG, European Crohn’s and Colitis Organization (ECCO), International Organization for Inflammatory Bowel Disease (IOIBD), and other groups were specifically reviewed and incorporated alongside prior topic specific systematic or evidence-based reviews.^4,9-12,17-23^ The literature review was synthesized into a summary statement with relevant references which was distributed to all panelists before the first round of ratings.

### Clinical Scenarios

The survey was divided into 2 sections broadly related to steroid management and maintenance therapy, and variables assessed for the different clinical scenarios were selected based on their potential influence on decision making and/or patient outcomes. In section 1 (steroid management), we had two chapters related to in-hospital and post-discharge steroid dosing. In-hospital steroid dosing took into consideration different formulations (methylprednisolone or hydrocortisone), dosages, need for rescue medical therapy with infliximab or cyclosporine, and need for monitoring during transition to oral prednisone regimens prior to discharge. Post-discharge steroid dosing took into consideration the in-hospital steroid dose used, in-hospital response to steroids and/or need for rescue infliximab or cyclosporine, overall risk for adverse events, and outpatient prednisone dosage. In total we had 43 statements during the first round of voting, and 45 statements in the second round of voting that were rated by panelists. In section 2 (maintenance therapy), we had 2 chapters related to: hospital discharge criteria and post-discharge timing of assessment of disease activity, and post-discharge optimization or initiation of biologics or small molecule therapies. Post-discharge biologic or small molecule therapy scenarios took into consideration in-hospital response to steroids and/or need for rescue infliximab or cyclosporine, overall risk for adverse events, and exposure to biologics or small molecule therapies prior to hospitalization. This was assessed separately for anti-TNF agents, vedolizumab, ustekinumab, ozanimod, and tofacitinib. In total we had 50 statements during the first round of voting, and 49 statements in the second round of voting that were rated by panelists. In total, there were 94 final statements voted on by panelists during the second round of voting.

### Definitions

RAND panels require specific definitions and assumptions to manage the impractical number of scenarios if we tried to address every possible combination of variables. These definitions and assumptions are made clear to the panel before any voting takes place. We defined moderate-severe UC flares as a full Mayo score of 6-12 with all sub-scores 2-3. Partial response was defined as at least a 1-point reduction in rectal bleeding and stool frequency Mayo sub-scores, and remission was defined as resolution of rectal bleeding (Mayo rectal bleeding sub-score of 0) and at least a 1-point reduction in the Mayo stool frequency sub-score. Overall risk of adverse events was categorized as low or high, with high-risk being defined broadly as age > 65 years or presence of comorbidities increasing the risks associated with immune suppression.

### Assumptions

We made several assumptions for this panel that included:

- Hospitalization is for a moderate-severe UC flare, which is more broadly encompassing of UC flare related hospitalizations as compared to traditional acute severe UC criteria
- There are no complicating secondary factors (e.g., infection with *Clostridioides difficile* or Cytomegalovirus)
- Post-discharge care time we are addressing is 12 weeks post-discharge
- Rescue therapy considered is either intravenous (IV) steroids alone, IV steroids + infliximab, or IV steroids + cyclosporine
- All patients being discharged without surgery had either a partial response or remission to rescue medical therapy
- Patients will be discharged on a prednisone dose equivalent or lower than the methylprednisolone or hydrocortisone dose used in the hospital
- If a patient had a partial response or remission to infliximab rescue therapy in the hospital, it will be continued with optimized dosing based on therapeutic drug monitoring (TDM) after discharge
- If a patient had partial response or remission to cyclosporine rescue therapy in the hospital, they would transition to a biologic or small molecule for UC maintenance after discharge
- Outpatient maintenance biologic/small molecule will be optimized based on societal guidelines for TDM target concentrations^24^
- The use of combination therapy (an immunomodulator with the biologic) is based on the provider and patient discussion balancing benefits, risks, and patient preferences
- Clinic appointment post-discharge could be in-person or virtual

### Appropriateness Panel and Rating

The panel included members of the ongoing National Institute of Diabetes and Digestive and Kidney Diseases (NIDDK) sponsored U34 planning grant for hyperbaric oxygen therapy in hospitalized ulcerative colitis (HBOT-UC). After receiving the literature summary, panelists confidentially rated the statements through a web-based questionnaire (Google Forms) and convened 2 weeks later at a moderated virtual session. During this meeting statements with median scores in the uncertain range or those with a high disagreement index were discussed in detail, and clarifications were made regarding statement phrasing or questions surrounding interpretation of statements. Revisions to statements were made or new statements were added to address these clarifications where needed, and the panelists then re-rated each statement confidentially. The goal of this panel was not to achieve consensus but to rate level of appropriateness of each statement.

### Analysis

Appropriateness was rated on a scale of 1 to 9 for each statement (1-3, inappropriate; 4-6, uncertain; and 7-9, appropriate). Median scores were calculated and rounded up, so that a median score of 3.5 was rated as uncertain whereas a median score of 6.5 was rated as appropriate. To quantify the level of agreement, a RAND disagreement index (DI) was calculated for each statement using a standard published equation.^**13**^ A DI greater than or equal to 1.0 indicates extreme variation whereas DI values less than 1.0 reflect general agreement. The DI expresses the spread of responses and is calculated using a previously described approach and the following formula^**13**^:

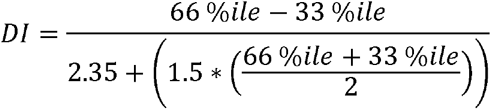

Statements in which ratings met criteria for disagreement were rated “uncertain” regardless of the median appropriateness score.

## RESULTS

A total of 16 panelists voted in the first round, of which 14 attended the moderated virtual session. These 14 panelists then re-voted in the second round and the 2 panelists who could not attend the moderated session did not vote in the second round based on RAND methodology. Detailed results are presented in **Supplementary Table 1**.

### In-hospital steroid dosing strategies

Panelists rated 60mg methylprednisolone IV every 24 hours, and its equivalent hydrocortisone dose of 100mg IV three times a day, as “appropriate” with a median rating of 9 for each and low interquartile ranges (IQR; 0 and 1) and DI scores (0 and -0.34). Panelists also rated 40mg methylprednisolone IV every 24 hours as “appropriate” with a median rating of 7 but a wide IQR (3.75) and DI (−23.06). They rated the equivalent hydrocortisone dose of 100mg IV twice daily as “uncertain”. (**Table 1**) During the moderated virtual session panelists discussed needing to weigh the relative safety of higher doses of methylprednisolone or hydrocortisone against the need to achieve rapid improvements in disease activity and avoid in-hospital progression or toxicity requiring emergent colectomy. Safety or intolerance of higher dose steroids was a primary consideration for some panelists when voting for 40mg methylprednisolone, and a new statement was added during the second round of voting regarding the use of 40mg methylprednisolone IV every 24 hours if patients are intolerant to higher steroid doses which was rated as “appropriate” with a median rating of 9, IQR of 1, and DI of -0.23. Hydrocortisone 100mg IV twice daily was still rated as “uncertain” in the new statement.

**Table 1:** Recommendations on inpatient steroid dosing for acute moderate-severe disease flares.

| **Statement** | **Recommendation** |
| --- | --- |
| It is appropriate to use 60mg methylprednisolone intravenous every 24 hours for the in-hospital management of a moderate-severe UC flare. | Appropriate |
| It is appropriate to use 100mg hydrocortisone intravenous three times a day for the in-hospital management of a moderate-severe UC flare. | Appropriate |
| It is appropriate to use 40mg methylprednisolone intravenous every 24 hours for the in-hospital management of a moderate-severe UC flare. | Appropriate |
| It is appropriate to use 40mg methylprednisolone intravenous every 24 hours for the in-hospital management of a moderate-severe UC flare if patients are intolerant to higher steroid doses. | Appropriate |
| It is appropriate to use 100mg hydrocortisone intravenous two times a day for the in-hospital management of a moderate-severe UC flare. | Uncertain |
| It is appropriate to use 100mg hydrocortisone intravenous two times a day for the in-hospital management of a moderate-severe UC flare if patients are intolerant to higher steroid doses. | Uncertain |
| It is appropriate to continue intravenous steroids if transitioning to second line medical therapy (infliximab or cyclosporine) in the hospital after a partial response to IV steroids. | Appropriate |
| It is appropriate to continue intravenous steroids if transitioning to second line medical therapy (infliximab or cyclosporine) in the hospital after complete non-response to IV steroids, | Uncertain |
| It is appropriate to observe patients for 24 hours on oral prednisone prior to discharge. | Appropriate |

Panelists rated the continued use of IV steroids when starting in-hospital infliximab or cyclosporine as “appropriate” if a partial response to IV steroids was observed prior to initiation of rescue therapy, however, the continued use of IV steroids when starting in-hospital infliximab or cyclosporine was rated as “uncertain” if patients had complete non-response to IV steroids prior to starting these rescue agents. The discussion around this statement focused on the fact that continued use of steroids in addition to a second immunosuppressive agent was felt to add unnecessary safety risks given the complete non-response, however, some felt that taking away steroids during a high-risk period might allow for worsening of disease activity and progression to colectomy. Therefore, although the median rating was 7, this statement of continuing IV steroids when starting cyclosporine or infliximab despite complete non-response to IV steroids, was “uncertain” due to a high DI (2.35).

### Outpatient prednisone dosing strategies after discharge

Panelists rated it “appropriate” that patients should be observed for 24 hours on oral prednisone prior to discharge. The use of 60mg of oral prednisone once daily was rated as “uncertain” or “inappropriate” across scenarios. (**Figure 1**) The use of 40mg of oral prednisone once daily was rated as “appropriate” across all scenarios irrespective of whether patients had received 60mg or 40mg of IV methylprednisolone (or equivalent hydrocortisone dosing) in-hospital. (**Figure 1, Supplementary Table 1**) The panelists specifically reviewed that the equivalent oral prednisone dose of 60mg IV methylprednisolone was 75mg, and both 60mg and 40mg of oral prednisone was a 20% to nearly 50% dose reduction on discharge. Several panelists felt that given the outpatient steroid dose was going to be used for an extended period (> 2 weeks) possibly alongside other immunosuppressive agents started in-hospital or shortly after discharge, safety considerations for higher dose steroids needed to be factored into discharge dosing and outpatient steroid management plans even among those receiving 60mg of methylprednisolone inpatient. Some panelists did not that this would need to be balanced against the risk of relapse and re-admission, particularly among those reducing from 60mg IV methylprednisolone to 40mg oral prednisone, and emphasized that close follow-up and monitoring would be needed if this were done.

**Figure 1:**
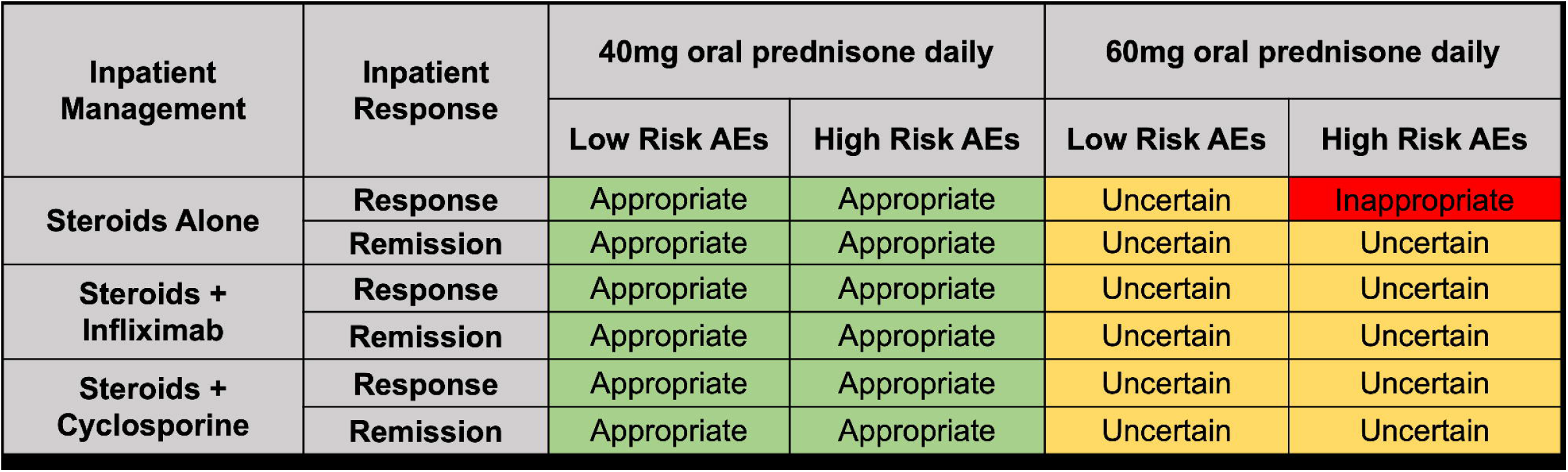
Recommendations on outpatient steroid dosing after discharge for acute moderate-severe disease flares where 60mg IV methylprednisolone daily or 100mg IV hydrocortisone three times daily used. AE: Adverse events. The definition of high risk for AEs was age > 65 years or presence of comorbidities increasing the risks associated with immune suppression.

**Figure 2:**
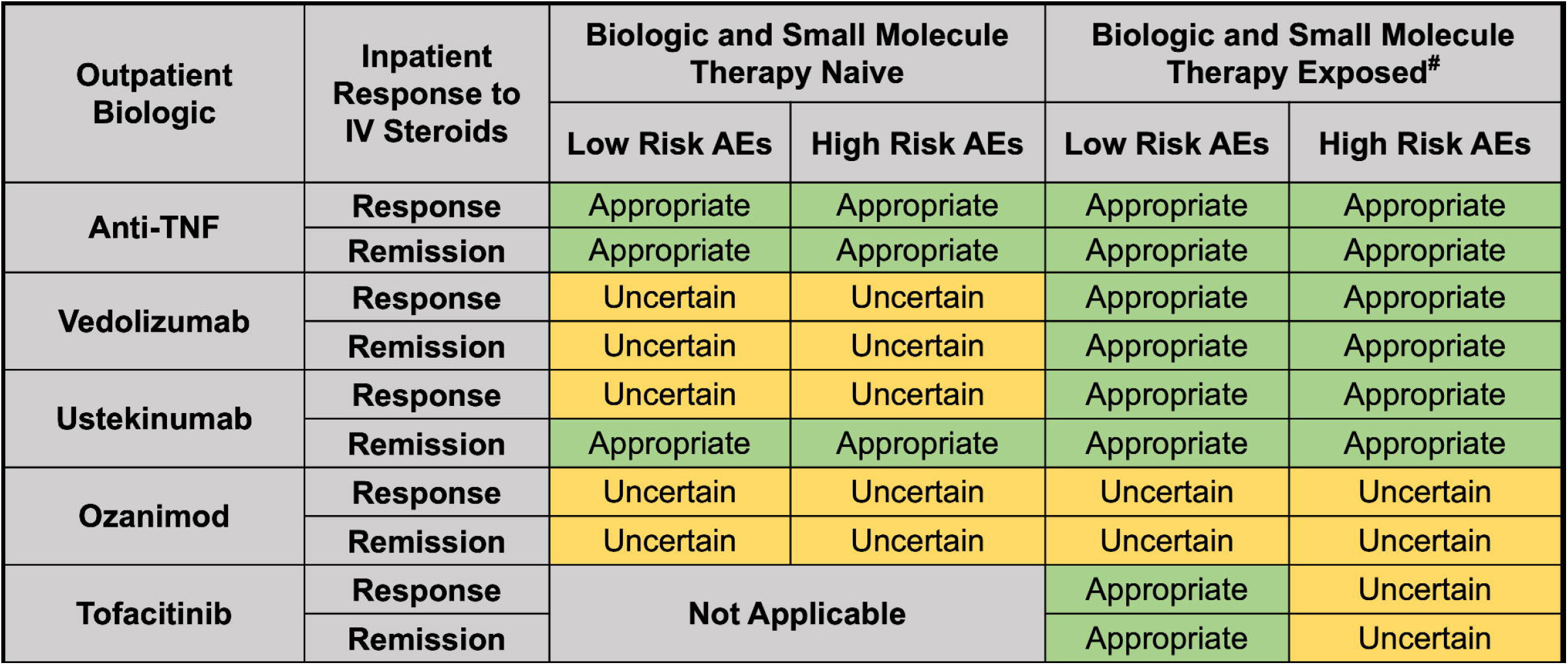
Outpatient biologic or small molecule initiation after discharge for acute moderate-severe UC. ^#^Biologic and small molecule exposure did not include exposure to within class agents. For anti-TNF therapy, we did not assess the scenario of starting an anti-TNF therapy post-discharge in a patient who had previously been exposed to an anti-TNF agent prior to hospitalization. AE: Adverse events. The definition of high risk for AEs was age > 65 years or presence of comorbidities increasing the risks associated with immune suppression.

### Discharge criteria and timing of follow-up for disease activity assessments

Panelists rated it “appropriate” to discharge patients from the hospital once they have achieved resolution of rectal bleeding. (**Table 2**) During the first round, discharging patients who achieved normalization of stool frequency and form but with persistent rectal bleeding was rated as “uncertain”. During the moderated session panelists felt it would be implausible that a patient would have persistent significant rectal bleeding but normalization of stool frequency and form, and the presence of mild bleeding (Mayo rectal bleeding score of 1) in this setting was not felt to be enough to keep a patient in the hospital when bowel movement frequency and form had normalized. During the moderated discussion a new question was added for the second round, and panelists voted that it was “appropriate” to discharge a patient when either rectal bleeding had resolved, or stool frequency had returned to baseline frequency and form. After discharge, panelists rated it “appropriate” to follow-up patients within 2 weeks for a clinic appointment and within 4-6 months for a sigmoidoscopy or colonoscopy. It was rated “inappropriate” to wait as long as 8 weeks after discharge to have the first follow-up clinic appointment.

**Table 2:** Recommendations on discharge and follow-up criteria after hospitalization for acute-moderate-severe disease flares.

| Statement | Recommendation |
| --- | --- |
| It is appropriate to discharge patients from the hospital when visible blood in their stool has resolved and they are having stools at their baseline frequency and form. | Appropriate |
| It is appropriate to discharge patients from the hospital when visible blood in their stool has resolved but stool frequency is still above their baseline. | Appropriate |
| It is appropriate to discharge patients from the hospital when either the visible blood in their stool has resolved or their stools are at their baseline frequency and form. | Appropriate |
| It is appropriate to discharge patients from the hospital when visible blood is still present in their stool and stool frequency is still above their baseline. | Inappropriate |
| It is appropriate to follow-up patients with a clinic appointment after discharge within 2 weeks. | Appropriate |
| It is appropriate to follow-up patients with a clinic appointment after discharge within 4 weeks. | Uncertain |
| It is appropriate to follow-up patients with a clinic appointment after discharge within 8 weeks. | Inappropriate |
| It is appropriate to follow-up patients with a sigmoidoscopy or colonoscopy after discharge within 2 months. | Uncertain |
| It is appropriate to follow-up patients with a sigmoidoscopy or colonoscopy after discharge within 4-6 months. | Appropriate |

### Optimization of biologics or small molecules used prior to hospitalization versus switching

Panelists rated it “appropriate” to continue and optimize the biologic or small molecule therapy patients were on prior to hospitalization if the patient achieved remission with IV steroids alone in the hospital. For patients achieving only a partial response to IV steroids in the hospital, continued use of the prior biologic or small molecule was rated as “uncertain” due to a high DI. During the moderated session panelists commented that these scenarios of being discharged after remission or a partial response to IV steroids still needed to be addressed on a case-by-case basis after taking into consideration number of prior biologic failures, duration of pre-admission treatment and potential for dose optimization of the biologic or small molecule therapy the patient was on prior to hospitalization, risk for disease progression after discharge particularly for those who had not achieved remission in-hospital, and rationale for discharging with only partial response to IV steroids with regards to planned outpatient therapy. Panelists rated it “appropriate” to switch to an alternative biologic or small molecule agent after discharge for patients achieving remission or partial response to IV steroids alone in the hospital. (**Supplementary Table 1**)

### Post-discharge initiation of a new biologic or small molecule

When considering which biologic or small molecule to start post-discharge among patients treated with steroids alone in the hospital, anti-TNF therapy was rated “appropriate” among anti-TNF naïve patients across all scenarios including if they had been exposed to biologics other than anti-TNF agents or small molecules prior to hospitalization. Vedolizumab and ustekinumab were rated as “appropriate” for patients with prior biologic or small molecule exposure, and tofacitinib was rated as “appropriate” for patients with prior anti-TNF exposure and a low risk of adverse events. Uncertainty existed when considering vedolizumab or ustekinumab in biologic or small molecule naïve patients, ozanimod in the post-discharge setting, and tofacitinib for patients with prior anti-TNF exposure and a high risk of adverse events. During the moderated discussion these “uncertain” ratings for vedolizumab and ustekinumab in biologic and small molecule naïve patient scenarios was noted to be largely driven by the fact that anti-TNF therapy is a treatment option in these scenarios and would be favored. The use of ozanimod after discharge was rated as “uncertain” across all scenarios, and this was attributed solely to it being a newer agent. The uncertainty for tofacitinib was due to the FDA labeling changes and long-term safety observations in rheumatoid arthritis studies, but it was noted that we had not differentiated between risks specific to tofacitinib such as major adverse cardiac events, or general risk to all immunosuppressives, which would be important for patient-specific application of these recommendations.

## DISCUSSION

UC patients hospitalized for an acute flare represent one of our highest risk patient populations, but limited guidance exists on the appropriate treatment and monitoring strategy to optimize health outcomes. Through a RAND Appropriateness Panel, we provide guidance on the appropriate strategy for steroid dosing, discharge criteria, follow-up assessments, and outpatient biologic or small molecule therapy post-discharge. We have made several key observations that will help inform optimal care for these patients.

First, in-hospital steroid dosing should be 60mg methylprednisolone IV every 24 hours or 100mg hydrocortisone IV three times daily (both are equivalent steroid doses), with 40mg methylprednisolone IV every 24 hours being used in patients who are known to be intolerant to higher doses of steroids. This aligns with ACG guidelines which recommend 60mg methylprednisolone IV every 24 hours or 100mg hydrocortisone IV three to four times daily.^9^ The AGA guidelines do not provide clear recommendations and allow for a choice between 40-60mg methylprednisolone IV every 24 hours for the management of acute flares.^10^ Although 40mg of methylprednisolone was rated as appropriate by the panelists, there was a wide range in panelist voting and over one-third of panelists voted the appropriateness of this dose to be uncertain or inappropriate. During the moderated discussion, the panel felt that although higher doses of steroids are associated with an increased risk for adverse events, this was primarily in the outpatient setting over longer periods of use and the in-hospital risk of under-treating the disease outweighed the short-term risks of higher dose steroids. Therefore, a new statement was added and there was broad agreement that 40mg methylprednisolone IV every 24 hours was appropriate if patients were known to be intolerant to higher doses.

Second, there was broad agreement across all scenarios that 40mg of prednisone was an appropriate dose upon discharge even among patients using 60mg of methylprednisolone/300mg hydrocortisone daily in-hospital, which converts to 75mg of prednisone daily. The panel acknowledged this dose reduction during the moderated discussion, but still felt the 40mg of prednisone was appropriate during the second round of voting due to safety concerns with higher doses of outpatient prednisone which would result in prolonged tapering and longer periods of exposure to high-dose (> 20mg) steroids potentially alongside additional immunosuppressive agents. The prior clinical trial comparing 20mg to 40mg to 60mg of prednisone was discussed as this is often cited for why 40mg is appropriate in the outpatient setting.^18^ It was noted that this trial is very old with a small sample size for comparison (n=20 per arm), and the goals were to understand if doses > 20mg were superior rather than trying to directly compare 40mg to 60mg. Furthermore, this trial was not directly applicable to the post-discharge setting. The ACG guidelines recommend 40-60mg of prednisone for outpatient UC flares,^9^ and therefore do allow for a higher dose of prednisone in the outpatient setting. Some panelists felt that this higher dose may still be appropriate on a case-by-case basis particularly in patients who are low risk for adverse events and are being discharged after achieving partial response to inpatient IV steroids without the need for inpatient infliximab or cyclosporine. During the moderated discussion, panelists noted that an approach to avoiding prolonged exposure to higher doses with 60mg was to do a taper of 10mg weekly until the patient was at 40mg as a replacement for the 2 weeks at 40mg often done, and then continuing with traditional dose reductions used in clinical practice.

Third, there is no guidance on discharge criteria or post-discharge monitoring for UC patients hospitalized for acute flares. The panel felt it was appropriate to use resolution of rectal bleeding or normalization of stool frequency and form as criteria for discharge. The achievement of both normalization of rectal bleeding and normalization of bowel movement frequency and form was felt to be the most ideal discharge criteria, but panelists also felt it was appropriate to discharge patients if rectal bleeding had completely resolved even if stool frequency was above baseline form and frequency. It was felt unlikely that a scenario would arise where a patient achieved normalized stool frequency and form but with persistent moderate-severe rectal bleeding (Mayo rectal bleeding sub-score 2 or 3), and therefore it was felt to be appropriate for patients to be discharged if stool frequency and form had normalized and only mild rectal bleeding was present. This discussion led to the modified question for either resolution of rectal bleeding or normalization in stool frequency and form, and providers should note that this does not apply to the anticipated rare proportion of patients who have normalized stool frequency and form but persistent moderate-severe rectal bleeding.

The total daily steroid dose reduction from IV to PO steroid dosing was a primary reason why it was felt to be appropriate to observe patients on oral prednisone for 24 hours prior to discharge to help avoid acute worsening and return to the emergency department. Once discharged, the panel felt it was appropriate to follow-up patients clinically within 2 weeks to ensure optimal care coordination, treatment optimization, and monitoring for disease worsening particularly when considering the dose reduction in prednisone upon discharge. The IOIBD has stated that endoscopic healing is a long-term target and recommended that in a treat-to-target monitoring strategy UC patients should be assessed with lower endoscopy every 4-6 months until remission is achieved.^11,23^ The post-discharge setting was not specifically discussed by IOIBD, however, this RAND panel agreed on this timeline as being appropriate after discharge for an acute flare.

Finally, the initiation of a new biologic or small molecule in the post-discharge setting was reviewed. These results should be prefaced by stating that all biologics or small molecules reviewed are FDA approved for use in outpatient moderate-severe UC patients. Therefore, a rating of “uncertain” does not imply a lack of efficacy but rather a degree of uncertainty as to whether the biologic or small molecule is most ideal for that given scenario. Broad agreement was present that anti-TNF therapy was appropriate across all scenarios for patients naïve to anti-TNF therapy prior to hospitalization given the evidence base for infliximab. This point factored into panelists voting for other biologics as well. Among biologic or small molecule exposed patients both vedolizumab and ustekinumab were rated as appropriate, but less certainty existed on their use among anti-TNF naïve patients because panelists felt anti-TNF therapy had the greatest evidence for disease modification. In anti-TNF exposed patients, tofacitinib was felt to also be appropriate if they were at low risk for adverse events and panelists commented that it may be considered in those with a high risk of adverse events on a case-by-case basis. Although the use of ozanimod was rated as uncertain by panelists across all scenarios, it was noted and discussed that this was not due to lack of efficacy but rather lack of provider experience with this drug. Ozanimod is FDA approved for use in outpatient moderate-severe UC patients and therefore remains a viable treatment with proven efficacy in phase 3 clinical trial programs.^25^ Ultimately, decisions on initiation of biologics and small molecules post-discharge will need to take into consideration individual patient demographics, prognostic factors such as C-reactive protein and albumin, overall risk of disease or treatment complications, and therapy specific personalized decision support tools.

Although we were able to provide recommendations for multiple scenarios, some areas of uncertainty remained. A notable point of discussion was whether providers should continue IV steroids when transitioning to infliximab or cyclosporine if patients had complete non-response to IV steroids prior to this transition. The primary disagreement for this scenario was around the added risk of dual immunosuppression with unclear clinical benefit and variability in defining non-response. Panelists noted that providers will need to consider the totality of data and even small changes in C-reactive protein might suggest some benefit in continued use to avoid acute worsening and need for emergent colectomy, but all panelists agreed that this scenario had very limited evidence or guidance on the appropriateness of continuing versus stopping IV steroids.

Our study has multiple strengths including the importance of the topics being addressed, the application of the well validated and robust RAND methodology, and the consistency in rating of panelists across similarly clustered concepts or themes for scenarios. There are some important limitations worth noting when interpreting these results. As most RAND panels are performed because clinical guidance is still needed despite the lack of data, we also lack evidence to guide many of the scenarios. In the setting of uncertainty and paucity of data, voting on hypothetical patient scenarios in the setting of a RAND panel is better than no recommendation, but we need to recognize the variability of real-life patients and logistics. Along these lines we could not review every possible scenario due to the multiplicative nature of variables being added and subsequently creating hundreds to thousands of possible scenarios for voting. Therefore, while we have attempted to address the scenarios felt to be of highest clinical utility, we recognize that some patient scenarios are not accounted for in this study. Although being able to address clinical scenarios with limited evidence base is a primary strength of the RAND methodology, it will be important to obtain evidence going forward to better inform these decisions. Panelists were instructed to not consider cost or patient preferences; however, the post-discharge use of biologics or small molecules will be influenced by payors, out-of-pocket expenses, and patient perceptions, and decisions will need to be made on an individual basis with shared decision-making approaches. Finally, we have not considered the emerging approach of tofacitinib rescue therapy in-hospital. As continued evidence is generated for its use in this setting, providers will need to be particularly thoughtful about this scenario when considering the continued use of IV steroids.

In conclusion, we provide recommendations for the in-hospital and post-discharge care of UC patients hospitalized for acute flares. These recommendations help to clarify appropriate steroid dosing, discharge criteria, patient monitoring, and post-discharge use of biologics or small molecules. These recommendations cannot replace clinical judgement, and should still be considered in the context of available societal guidelines, but it is our hope that this evidence will create more uniform care for this high-risk population.

## Data Availability

All data produced in the present work are contained in the manuscript

