## Supplementary Table 1 for "Appropriate management of steroids and discharge planning during and after hospital admission for moderate-severe ulcerative colitis"

**Supplementary Table 1: RAND Appropriateness Panel results and recommendations on steroid management, discharge criteria, and post-discharge care for ulcerative colitis patients hospitalized for acute moderate-severe disease flares**

|  | **Pre-Survey (n=16): 93 Statements (54 Appropriate, 37 Uncertain, 2 Inappropriate)** | | | | | | |  | **Post-Survey (n=14): 94 Statements (58 Appropriate, 33 Uncertain, 3 Inappropriate)** | | | | | | |
| --- | --- | --- | --- | --- | --- | --- | --- | --- | --- | --- | --- | --- | --- | --- | --- |
| **Statement** | **Median** | **IQR** | **Mean** | **SD** | **Median Category** | **DI** | **FINAL CATEGORY** |  | **Median** | **IQR** | **Mean** | **SD** | **Median Category** | **DI** | **FINAL CATEGORY** |
| **STEROID MANAGEMENT** |  |  |  |  |  |  |  |  |  |  |  |  |  |  |  |
| It is appropriate to use 𝟲𝟬𝗺𝗴 solumedrol intravenous every 24 hours for the in-hospital management of a moderate-severe UC flare. | 8.5 | 2 | 7.81 | 2.01 | Appropriate | -0.37 | Appropriate |  | 9 | 0 | 8.1 | 2.3 | Appropriate | 0.00 | Appropriate |
| It is appropriate to use 𝟰𝟬𝗺𝗴 solumedrol intravenous every 24 hours for the in-hospital management of a moderate-severe UC flare. | 7.5 | 2.25 | 7.19 | 2.29 | Appropriate | -0.93 | Appropriate |  | 7 | 3.75 | 6.2 | 2.5 | Appropriate | -23.06 | Appropriate |
| NEW STATEMENT: It is appropriate to use 𝟰𝟬𝗺𝗴 solumedrol intravenous every 24 hours for the in-hospital management of a moderate-severe UC flare if patients are intolerant to higher steroid doses. | --- |  |  |  |  |  |  |  | 9 | 1 | 8.4 | 1.0 | Appropriate | -0.23 | Appropriate |
| It is appropriate to use 100mg hydrocortisone intravenous 𝗧𝗛𝗥𝗘𝗘 times a day for the in-hospital management of a moderate-severe UC flare. | 8 | 1.25 | 7.50 | 1.71 | Appropriate | -0.71 | Appropriate |  | 9 | 1 | 8.4 | 0.8 | Appropriate | -0.34 | Appropriate |
| It is appropriate to use 100mg hydrocortisone intravenous 𝗧𝗪𝗢 times a day for the in-hospital management of a moderate-severe UC flare. | 5.5 | 3 | 5.13 | 2.47 | Uncertain | 0.85 | Uncertain |  | 4 | 1.75 | 4.4 | 1.5 | Uncertain | 0.32 | Uncertain |
| NEW STATEMENT: It is appropriate to use 100mg hydrocortisone intravenous 𝗧𝗪𝗢 times a day for the in-hospital management of a moderate-severe UC flare if patients are intolerant to higher steroid doses. | --- |  |  |  |  |  |  |  | 7 | 1.75 | 6.5 | 2.2 | Appropriate | 10.00 | Uncertain |
| It is appropriate to continue intravenous steroids if transitioning to second line medical therapy (infliximab or cyclosporine) in the hospital after a 𝘱𝘢𝘳𝘵𝘪𝘢𝘭 𝘳𝘦𝘴𝘱𝘰𝘯𝘴𝘦 to IV steroids. | 8 | 2 | 7.56 | 1.50 | Appropriate | -0.71 | Appropriate |  | 8 | 0.75 | 7.9 | 1.1 | Appropriate | 0.00 | Appropriate |
| It is appropriate to continue intravenous steroids if transitioning to second line medical therapy (infliximab or cyclosporine) in the hospital after 𝘤𝘰𝘮𝘱𝘭𝘦𝘵𝘦 𝘯𝘰𝘯-𝘳𝘦𝘴𝘱𝘰𝘯𝘴𝘦 to IV steroids, | 6 | 2.5 | 5.75 | 2.41 | Uncertain | 2.05 | Uncertain |  | 6.5 | 2 | 5.6 | 2.4 | Appropriate | 2.35 | Uncertain |
| It is appropriate to observe patients for 24 hours on oral prednisone prior to discharge. | 8 | 2 | 7.81 | 1.05 | Appropriate | -0.71 | Appropriate |  | 8 | 1.75 | 8.1 | 0.9 | Appropriate | -0.34 | Appropriate |
| **1. It is appropriate to discharge a patient on 60mg of prednisone daily if they received the equivalent of 60mg of solumedrol (or 100mg hydrocortisone TID) in the hospital if…** | | | | | | | | | | | | | | | |
| they have a low risk for AEs, and the patient had a 𝘱𝘢𝘳𝘵𝘪𝘢𝘭 𝘳𝘦𝘴𝘱𝘰𝘯𝘴𝘦 with IV steroid rescue therapy. | 5.5 | 5 | 5.38 | 2.68 | Uncertain | 1.70 | Uncertain |  | 6.5 | 3 | 5.6 | 2.3 | Appropriate | 1.96 | Uncertain |
| they have a HIGH risk for AEs, and the patient had a partial response with IV steroid rescue therapy. | 3.5 | 4.5 | 4.06 | 2.67 | Uncertain | 0.99 | Uncertain |  | 3 | 3.5 | 3.3 | 2.1 | Inappropriate | 0.24 | Inappropriate |
| they have a low risk for AEs, and the patient 𝘢𝘤𝘩𝘪𝘦𝘷𝘦𝘥 𝘳𝘦𝘮𝘪𝘴𝘴𝘪𝘰𝘯 with IV steroid rescue therapy. | 7 | 4.25 | 6.25 | 2.41 | Appropriate | -3.65 | Appropriate |  | 7 | 2.75 | 5.9 | 2.4 | Appropriate | 2.70 | Uncertain |
| they have a HIGH risk for AEs, and the patient achieved remission with IV steroid rescue therapy. | 5 | 5 | 4.63 | 2.53 | Uncertain | 0.97 | Uncertain |  | 4.5 | 3 | 4.1 | 2.0 | Uncertain | 0.76 | Uncertain |
| they have a low risk for AEs, and the patient had a 𝘱𝘢𝘳𝘵𝘪𝘢𝘭 𝘳𝘦𝘴𝘱𝘰𝘯𝘴𝘦 with IV steroid + 𝗶𝗻𝗳𝗹𝗶𝘅𝗶𝗺𝗮𝗯 rescue therapy. | 6.5 | 4.25 | 5.88 | 2.60 | Appropriate | 2.31 | Uncertain |  | 6 | 4.5 | 5.3 | 2.9 | Uncertain | 1.35 | Uncertain |
| they have a HIGH risk for AEs, and the patient had a partial response with IV steroid + infliximab rescue therapy. | 4 | 4.25 | 4.50 | 2.50 | Uncertain | 0.52 | Uncertain |  | 3.5 | 3.5 | 3.8 | 2.2 | Uncertain | 0.38 | Uncertain |
| they have a low risk for AEs, and the patient 𝘢𝘤𝘩𝘪𝘦𝘷𝘦𝘥 𝘳𝘦𝘮𝘪𝘴𝘴𝘪𝘰𝘯 with IV steroid + infliximab rescue therapy. | 6 | 4.5 | 5.25 | 2.82 | Uncertain | 1.86 | Uncertain |  | 6 | 4.5 | 5.4 | 2.9 | Uncertain | 1.88 | Uncertain |
| they have a HIGH risk for AEs, and the patient achieved remission with IV steroid + infliximab rescue therapy. | 4 | 3.5 | 4.19 | 2.46 | Uncertain | 0.48 | Uncertain |  | 4 | 2.5 | 4.1 | 2.4 | Uncertain | 0.22 | Uncertain |
| they have a low risk for AEs, and the patient had a 𝘱𝘢𝘳𝘵𝘪𝘢𝘭 𝘳𝘦𝘴𝘱𝘰𝘯𝘴𝘦 with IV steroid + 𝗰𝘆𝗰𝗹𝗼𝘀𝗽𝗼𝗿𝗶𝗻𝗲 rescue therapy. | 6.5 | 4.25 | 5.44 | 2.80 | Appropriate | 1.70 | Uncertain |  | 7 | 4.75 | 5.9 | 2.9 | Appropriate | 4.11 | Uncertain |
| they have a HIGH risk for AEs, and the patient had a partial response with IV steroid + cyclosporine rescue therapy. | 4 | 5 | 4.50 | 2.73 | Uncertain | 0.92 | Uncertain |  | 4 | 4.75 | 4.4 | 2.6 | Uncertain | 0.80 | Uncertain |
| they have a low risk for AEs, and the patient 𝘢𝘤𝘩𝘪𝘦𝘷𝘦𝘥 𝘳𝘦𝘮𝘪𝘴𝘴𝘪𝘰𝘯 with IV steroid + cyclosporine rescue therapy. | 6 | 5.25 | 5.06 | 2.91 | Uncertain | 1.70 | Uncertain |  | 6 | 3.5 | 5.3 | 2.6 | Uncertain | 2.35 | Uncertain |
| they have a HIGH risk for AEs, and the patient achieved remission with IV steroid + cyclosporine rescue therapy. | 4 | 5 | 4.19 | 2.56 | Uncertain | 0.49 | Uncertain |  | 4 | 2.75 | 4.1 | 2.4 | Uncertain | 0.38 | Uncertain |
| **2. It is appropriate to discharge a patient on 40mg of prednisone daily if they received the equivalent of 60mg of solumedrol (or 100mg hydrocortisone TID) in the hospital if…** | | | | | | | | | | | | | | | |
| they have a low risk for AEs, and the patient had a 𝘱𝘢𝘳𝘵𝘪𝘢𝘭 𝘳𝘦𝘴𝘱𝘰𝘯𝘴𝘦 with IV steroid rescue therapy. | 7 | 2.25 | 6.81 | 2.17 | Appropriate | -3.08 | Appropriate |  | 7 | 3 | 6.9 | 2.3 | Appropriate | -1.76 | Appropriate |
| they have a HIGH risk for AEs, and the patient had a partial response with IV steroid rescue therapy. | 7 | 2.5 | 5.88 | 2.50 | Appropriate | 2.35 | Uncertain |  | 7 | 0.75 | 6.4 | 1.7 | Appropriate | 0.00 | Appropriate |
| they have a low risk for AEs, and the patient 𝘢𝘤𝘩𝘪𝘦𝘷𝘦𝘥 𝘳𝘦𝘮𝘪𝘴𝘴𝘪𝘰𝘯 with IV steroid rescue therapy. | 8.5 | 2 | 7.81 | 2.01 | Appropriate | -0.37 | Appropriate |  | 9 | 1.5 | 8.4 | 0.9 | Appropriate | 0.00 | Appropriate |
| they have a HIGH risk for AEs, and the patient achieved remission with IV steroid rescue therapy. | 7 | 1.25 | 6.88 | 2.47 | Appropriate | -0.71 | Appropriate |  | 7 | 1 | 7.1 | 2.0 | Appropriate | -0.71 | Appropriate |
| they have a low risk for AEs, and the patient had a 𝘱𝘢𝘳𝘵𝘪𝘢𝘭 𝘳𝘦𝘴𝘱𝘰𝘯𝘴𝘦 with IV steroid + 𝗶𝗻𝗳𝗹𝗶𝘅𝗶𝗺𝗮𝗯 rescue therapy. | 7.5 | 1.25 | 7.19 | 2.01 | Appropriate | -0.71 | Appropriate |  | 7.5 | 1 | 7.1 | 2.0 | Appropriate | -0.71 | Appropriate |
| they have a HIGH risk for AEs, and the patient had a partial response with IV steroid + infliximab rescue therapy. | 7 | 1.25 | 6.19 | 2.26 | Appropriate | -0.08 | Appropriate |  | 7 | 0 | 6.5 | 1.7 | Appropriate | 0.00 | Appropriate |
| they have a low risk for AEs, and the patient 𝘢𝘤𝘩𝘪𝘦𝘷𝘦𝘥 𝘳𝘦𝘮𝘪𝘴𝘴𝘪𝘰𝘯 with IV steroid + infliximab rescue therapy. | 8 | 2 | 7.44 | 2.28 | Appropriate | -0.37 | Appropriate |  | 9 | 1 | 8.4 | 0.9 | Appropriate | -0.23 | Appropriate |
| they have a HIGH risk for AEs, and the patient achieved remission with IV steroid + infliximab rescue therapy. | 7 | 2.25 | 6.00 | 2.48 | Appropriate | 7.64 | Uncertain |  | 7 | 0.75 | 6.5 | 2.0 | Appropriate | 0.00 | Appropriate |
| they have a low risk for AEs, and the patient had a 𝘱𝘢𝘳𝘵𝘪𝘢𝘭 𝘳𝘦𝘴𝘱𝘰𝘯𝘴𝘦 with IV steroid + 𝗰𝘆𝗰𝗹𝗼𝘀𝗽𝗼𝗿𝗶𝗻𝗲 rescue therapy. | 7 | 2.5 | 6.75 | 2.18 | Appropriate | -0.74 | Appropriate |  | 7 | 1 | 7.1 | 2.0 | Appropriate | -0.71 | Appropriate |
| they have a HIGH risk for AEs, and the patient had a partial response with IV steroid + cyclosporine rescue therapy. | 7 | 2.5 | 6.13 | 2.47 | Appropriate | 7.64 | Uncertain |  | 7 | 0 | 6.4 | 1.9 | Appropriate | 0.00 | Appropriate |
| they have a low risk for AEs, and the patient 𝘢𝘤𝘩𝘪𝘦𝘷𝘦𝘥 𝘳𝘦𝘮𝘪𝘴𝘴𝘪𝘰𝘯 with IV steroid + cyclosporine rescue therapy. | 8 | 1.25 | 6.81 | 2.69 | Appropriate | -0.71 | Appropriate |  | 8 | 0.75 | 7.9 | 1.1 | Appropriate | 0.00 | Appropriate |
| they have a HIGH risk for AEs, and the patient achieved remission with IV steroid + cyclosporine rescue therapy. | 7 | 1.5 | 6.13 | 2.47 | Appropriate | 10.00 | Uncertain |  | 7 | 0 | 6.7 | 1.8 | Appropriate | 0.00 | Appropriate |
| **3. It is appropriate to discharge a patient on 40mg of prednisone daily if they received the equivalent of 40 mg of solumedrol (or 100mg hydrocortisone BID) in the hospital if…** | | | | | | | | | | | | | | | |
| they have a low risk for AEs, and the patient had a 𝘱𝘢𝘳𝘵𝘪𝘢𝘭 𝘳𝘦𝘴𝘱𝘰𝘯𝘴𝘦 with IV steroid rescue therapy. | 8 | 2 | 7.69 | 1.54 | Appropriate | -0.34 | Appropriate |  | 8 | 2.5 | 7.1 | 2.3 | Appropriate | -0.44 | Appropriate |
| they have a HIGH risk for AEs, and the patient had a partial response with IV steroid rescue therapy. | 7 | 1.75 | 6.56 | 2.53 | Appropriate | -0.68 | Appropriate |  | 7 | 1 | 6.9 | 2.1 | Appropriate | -0.53 | Appropriate |
| they have a low risk for AEs, and the patient 𝘢𝘤𝘩𝘪𝘦𝘷𝘦𝘥 𝘳𝘦𝘮𝘪𝘴𝘴𝘪𝘰𝘯 with IV steroid rescue therapy. | 9 | 1 | 8.44 | 0.81 | Appropriate | -0.34 | Appropriate |  | 9 | 1 | 8.4 | 0.9 | Appropriate | -0.23 | Appropriate |
| they have a HIGH risk for AEs, and the patient achieved remission with IV steroid rescue therapy. | 7 | 2 | 7.38 | 1.93 | Appropriate | -0.71 | Appropriate |  | 7 | 1.75 | 7.6 | 1.0 | Appropriate | -0.53 | Appropriate |
| they have a low risk for AEs, and the patient had a 𝘱𝘢𝘳𝘵𝘪𝘢𝘭 𝘳𝘦𝘴𝘱𝘰𝘯𝘴𝘦 with IV steroid + 𝗶𝗻𝗳𝗹𝗶𝘅𝗶𝗺𝗮𝗯 rescue therapy. | 8 | 2 | 7.94 | 1.24 | Appropriate | -0.37 | Appropriate |  | 8 | 2 | 7.4 | 2.1 | Appropriate | -0.63 | Appropriate |
| they have a HIGH risk for AEs, and the patient had a partial response with IV steroid + infliximab rescue therapy. | 7 | 0.75 | 6.75 | 1.81 | Appropriate | 0.00 | Appropriate |  | 7 | 2.25 | 6.5 | 2.1 | Appropriate | 0.00 | Appropriate |
| they have a low risk for AEs, and the patient 𝘢𝘤𝘩𝘪𝘦𝘷𝘦𝘥 𝘳𝘦𝘮𝘪𝘴𝘴𝘪𝘰𝘯 with IV steroid + infliximab rescue therapy. | 8 | 2 | 7.69 | 1.78 | Appropriate | -0.34 | Appropriate |  | 8.5 | 1 | 8.2 | 1.0 | Appropriate | -0.34 | Appropriate |
| they have a HIGH risk for AEs, and the patient achieved remission with IV steroid + infliximab rescue therapy. | 7 | 1.5 | 6.88 | 2.06 | Appropriate | 0.00 | Appropriate |  | 7 | 0 | 7.1 | 1.1 | Appropriate | 0.00 | Appropriate |
| they have a low risk for AEs, and the patient had a 𝘱𝘢𝘳𝘵𝘪𝘢𝘭 𝘳𝘦𝘴𝘱𝘰𝘯𝘴𝘦 with IV steroid + 𝗰𝘆𝗰𝗹𝗼𝘀𝗽𝗼𝗿𝗶𝗻𝗲 rescue therapy. | 7.5 | 2 | 7.69 | 1.25 | Appropriate | -0.92 | Appropriate |  | 7 | 1.75 | 7.1 | 2.1 | Appropriate | -0.71 | Appropriate |
| they have a HIGH risk for AEs, and the patient had a partial response with IV steroid + cyclosporine rescue therapy. | 7 | 1.25 | 6.50 | 1.79 | Appropriate | -0.08 | Appropriate |  | 7 | 1 | 6.3 | 1.5 | Appropriate | -6.04 | Appropriate |
| they have a low risk for AEs, and the patient 𝘢𝘤𝘩𝘪𝘦𝘷𝘦𝘥 𝘳𝘦𝘮𝘪𝘴𝘴𝘪𝘰𝘯 with IV steroid + cyclosporine rescue therapy. | 8 | 2 | 7.44 | 1.93 | Appropriate | -0.71 | Appropriate |  | 8 | 1.75 | 8.1 | 0.8 | Appropriate | -0.22 | Appropriate |
| they have a HIGH risk for AEs, and the patient achieved remission with IV steroid + cyclosporine rescue therapy. | 7 | 1.25 | 6.63 | 2.03 | Appropriate | -0.08 | Appropriate |  | 7 | 0.75 | 7.1 | 1.0 | Appropriate | 0.00 | Appropriate |
| **MAINTENANCE THERAPY** |  |  |  |  |  |  |  |  |  |  |  |  |  |  |  |
| It is appropriate to discharge patients from the hospital when visible blood in their stool has resolved 𝗮𝗻𝗱 they are having stools at their baseline frequency and form. | 9 | 0 | 8.88 | 0.34 | Appropriate | 0.00 | Appropriate |  | 9 | 0 | 9.0 | 0.0 | Appropriate | 0.00 | Appropriate |
| It is appropriate to discharge patients from the hospital when visible blood in their stool has resolved 𝗯𝘂𝘁 stool frequency is still 𝗮𝗯𝗼𝘃𝗲 their baseline. | 8 | 2 | 7.50 | 1.93 | Appropriate | -0.71 | Appropriate |  | 8 | 1.75 | 7.4 | 2.0 | Appropriate | -0.71 | Appropriate |
| It is appropriate to discharge patients from the hospital when visible blood is 𝘀𝘁𝗶𝗹𝗹 𝗽𝗿𝗲𝘀𝗲𝗻𝘁 in their stool but they are having stools at their baseline frequency and form. | 6 | 4 | 5.94 | 2.26 | Uncertain | 13.88 | Uncertain |  |  |  |  |  |  |  |  |
| NEW STATEMENT: It is appropriate to discharge patients from the hospital when 𝗘𝗜𝗧𝗛𝗘𝗥 the visible blood in their stool has resolved 𝗢𝗥 their stools are at their baseline frequency and form. | --- |  |  |  |  |  |  |  | 7 | 1.75 | 6.7 | 1.9 | Appropriate | -0.53 | Appropriate |
| It is appropriate to discharge patients from the hospital when visible blood is still present in their stool and stool frequency is still 𝗮𝗯𝗼𝘃𝗲 their baseline. | 3 | 3.5 | 3.63 | 2.28 | Inappropriate | 0.62 | Inappropriate |  | 2.5 | 3.5 | 2.9 | 2.2 | Inappropriate | 0.29 | Inappropriate |
| It is appropriate to follow-up patients with a clinic appointment after discharge within 2 weeks. | 9 | 0.25 | 8.56 | 0.81 | Appropriate | 0.00 | Appropriate |  | 9 | 0 | 8.9 | 0.3 | Appropriate | 0.00 | Appropriate |
| It is appropriate to follow-up patients with a clinic appointment after discharge within 4 weeks. | 6.5 | 4.25 | 5.69 | 2.70 | Appropriate | 13.88 | Uncertain |  | 4 | 2.5 | 3.9 | 2.4 | Uncertain | 0.22 | Uncertain |
| It is appropriate to follow-up patients with a clinic appointment after discharge within 8 weeks. | 3 | 3 | 3.56 | 2.45 | Inappropriate | 0.35 | Inappropriate |  | 1 | 1.75 | 1.7 | 0.9 | Inappropriate | 0.13 | Inappropriate |
| It is appropriate to follow-up patients with a sigmoidoscopy or colonoscopy after discharge within 2 months. | 5 | 1.25 | 5.25 | 1.39 | Uncertain | 0.54 | Uncertain |  | 5 | 1.75 | 5.4 | 1.4 | Uncertain | 0.30 | Uncertain |
| It is appropriate to follow-up patients with a sigmoidoscopy or colonoscopy after discharge within 4 months. | 7 | 3 | 6.56 | 1.90 | Appropriate | 7.64 | Uncertain |  | --- |  |  |  |  |  |  |
| It is appropriate to follow-up patients with a sigmoidoscopy or colonoscopy after discharge within 6 months. | 8 | 1 | 8.25 | 0.77 | Appropriate | -0.34 | Appropriate |  | --- |  |  |  |  |  |  |
| CONSOLIDATED STATEMENT: It is appropriate to follow-up patients with a sigmoidoscopy or colonoscopy after discharge within 4-6 months. | --- |  |  |  |  |  |  |  | 9 | 1 | 8.5 | 0.7 | Appropriate | -0.34 | Appropriate |
| **1. Among patients treated with steroids alone in the hospital, on discharge it is appropriate to continue and optimize the biologic or small molecule inhibitor used prior to hospitalization if…** | | | | | | | | | | | | | | | |
| the patient had a 𝘱𝘢𝘳𝘵𝘪𝘢𝘭 𝘳𝘦𝘴𝘱𝘰𝘯𝘴𝘦 with IV steroid rescue therapy. | 6.5 | 2.25 | 5.94 | 2.29 | Appropriate | 2.35 | Uncertain |  | 7 | 2 | 6.1 | 2.1 | Appropriate | 3.42 | Uncertain |
| the patient 𝘢𝘤𝘩𝘪𝘦𝘷𝘦𝘥 𝘳𝘦𝘮𝘪𝘴𝘴𝘪𝘰𝘯 with IV steroid rescue therapy. | 7.5 | 3.25 | 6.94 | 2.26 | Appropriate | -0.77 | Appropriate |  | 8 | 2 | 7.7 | 1.6 | Appropriate | -0.72 | Appropriate |
| **2. Among patients treated with steroids alone in the hospital, on discharge it is appropriate to switch to a different biologic or small molecule inhibitor if…** | | | | | | | | | | | | | | | |
| the patient had a 𝘱𝘢𝘳𝘵𝘪𝘢𝘭 𝘳𝘦𝘴𝘱𝘰𝘯𝘴𝘦 with IV steroid rescue therapy. | 7 | 2.5 | 7.06 | 1.53 | Appropriate | -0.74 | Appropriate |  | 7 | 0.75 | 6.9 | 2.0 | Appropriate | 0.00 | Appropriate |
| the patient 𝘢𝘤𝘩𝘪𝘦𝘷𝘦𝘥 𝘳𝘦𝘮𝘪𝘴𝘴𝘪𝘰𝘯 with IV steroid rescue therapy. | 7 | 2.25 | 6.56 | 1.50 | Appropriate | 10.00 | Uncertain |  | 7 | 2 | 6.9 | 1.2 | Appropriate | -4.72 | Appropriate |
| **3. Among patients treated with steroids alone in the hospital, it is appropriate to start anti-TNF therapy after discharge if…** | | | | | | | | | | | | | | | |
| they have a low risk for AEs, the patient had a 𝘱𝘢𝘳𝘵𝘪𝘢𝘭 𝘳𝘦𝘴𝘱𝘰𝘯𝘴𝘦 with IV steroid rescue therapy, and is biologic/small molecule naïve prior to hospitalization. | 9 | 0.25 | 8.44 | 1.31 | Appropriate | 0.00 | Appropriate |  | 9 | 0 | 8.0 | 2.4 | Appropriate | 0.00 | Appropriate |
| they have a HIGH risk for AEs, the patient had a partial response with IV steroid rescue therapy, and is biologic/small molecule naïve prior to hospitalization. | 8 | 2 | 7.63 | 1.31 | Appropriate | -0.71 | Appropriate |  | 8 | 2 | 7.4 | 2.1 | Appropriate | -0.86 | Appropriate |
| they have a low risk for AEs, the patient had a partial response with IV steroid rescue therapy, and has been exposed to non-anti-TNF biologic/small molecule therapy prior to hospitalization. | 9 | 1 | 8.44 | 0.73 | Appropriate | -0.34 | Appropriate |  | 9 | 1.75 | 7.9 | 1.9 | Appropriate | -0.34 | Appropriate |
| they have a HIGH risk for AEs, the patient had a partial response with IV steroid rescue therapy, and has been exposed to non-anti-TNF biologic/small molecule therapy prior to hospitalization. | 8 | 1.25 | 7.63 | 1.26 | Appropriate | -0.71 | Appropriate |  | 8 | 1 | 7.2 | 2.1 | Appropriate | -0.44 | Appropriate |
| they have a low risk for AEs, the 𝘢𝘤𝘩𝘪𝘦𝘷𝘦𝘥 𝘳𝘦𝘮𝘪𝘴𝘴𝘪𝘰𝘯 with IV steroid rescue therapy, and is biologic/small molecule naïve prior to hospitalization. | 8 | 2 | 7.38 | 2.03 | Appropriate | -0.71 | Appropriate |  | 8.5 | 1.75 | 8.1 | 1.2 | Appropriate | -0.34 | Appropriate |
| they have a HIGH risk for AEs, the patient achieved remission with IV steroid rescue therapy, and is biologic/small molecule naïve prior to hospitalization. | 7 | 1.25 | 6.56 | 1.97 | Appropriate | -0.08 | Appropriate |  | 7 | 1.75 | 7.1 | 1.6 | Appropriate | -0.71 | Appropriate |
| they have a low risk for AEs, the patient achieved remission with IV steroid rescue therapy, and has been exposed to non-anti-TNF biologic/small molecule therapy prior to hospitalization. | 8 | 1.25 | 7.88 | 0.89 | Appropriate | -0.02 | Appropriate |  | 8 | 1 | 8.1 | 0.9 | Appropriate | -0.22 | Appropriate |
| they have a HIGH risk for AEs, the patient achieved remission with IV steroid rescue therapy, and has been exposed to non-anti-TNF biologic/small molecule therapy prior to hospitalization. | 8 | 1 | 7.44 | 1.26 | Appropriate | -0.71 | Appropriate |  | 8 | 2 | 7.9 | 1.2 | Appropriate | -0.63 | Appropriate |
| **4. Among patients treated with steroids alone in the hospital, it is appropriate to start vedolizumab after discharge if…** | | | | | | | | | | | | | | | |
| they have a low risk for AEs, the patient had a 𝘱𝘢𝘳𝘵𝘪𝘢𝘭 𝘳𝘦𝘴𝘱𝘰𝘯𝘴𝘦 with IV steroid rescue therapy, and is biologic/small molecule naïve prior to hospitalization. | 6 | 3.25 | 6.00 | 2.00 | Uncertain | 2.31 | Uncertain |  | 6 | 2 | 5.8 | 1.8 | Uncertain | 1.36 | Uncertain |
| they have a HIGH risk for AEs, the patient had a partial response with IV steroid rescue therapy, and is biologic/small molecule naïve prior to hospitalization. | 6 | 2.25 | 6.13 | 1.71 | Uncertain | 4.47 | Uncertain |  | 6 | 1.75 | 5.9 | 1.7 | Uncertain | 1.40 | Uncertain |
| they have a low risk for AEs, the patient had a partial response with IV steroid rescue therapy, and has been exposed to non-anti-integrin biologic/small molecule therapy prior to hospitalization. | 5 | 3 | 5.19 | 2.14 | Uncertain | 0.32 | Uncertain |  | 5 | 1.5 | 5.1 | 2.1 | Uncertain | 0.00 | Uncertain |
| they have a HIGH risk for AEs, the patient had a partial response with IV steroid rescue therapy, and has been exposed to non-anti-integrin biologic/small molecule therapy prior to hospitalization. | 6.5 | 3 | 6.44 | 1.82 | Appropriate | 2.35 | Uncertain |  | 5.5 | 2 | 5.8 | 2.1 | Uncertain | 2.35 | Uncertain |
| they have a low risk for AEs, the patient 𝘢𝘤𝘩𝘪𝘦𝘷𝘦𝘥 𝘳𝘦𝘮𝘪𝘴𝘴𝘪𝘰𝘯 with IV steroid rescue therapy, and is biologic/small molecule naïve prior to hospitalization. | 7.5 | 1 | 7.19 | 1.94 | Appropriate | -0.71 | Appropriate |  | 7.5 | 1 | 7.1 | 2.1 | Appropriate | -0.71 | Appropriate |
| they have a HIGH risk for AEs, the patient achieved remission with IV steroid rescue therapy, and is biologic/small molecule naïve prior to hospitalization. | 8 | 1 | 7.31 | 1.89 | Appropriate | -0.71 | Appropriate |  | 8 | 1 | 7.3 | 2.0 | Appropriate | -0.71 | Appropriate |
| they have a low risk for AEs, the patient achieved remission with IV steroid rescue therapy, and has been exposed to non-anti-integrin biologic/small molecule therapy prior to hospitalization. | 7 | 3 | 6.63 | 2.16 | Appropriate | -0.80 | Appropriate |  | 7 | 1.75 | 6.7 | 2.2 | Appropriate | -0.53 | Appropriate |
| they have a HIGH risk for AEs, the patient achieved remission with IV steroid rescue therapy, and has been exposed to non-anti-integrin biologic/small molecule therapy prior to hospitalization. | 7 | 2.25 | 6.75 | 1.91 | Appropriate | -3.08 | Appropriate |  | 7 | 2 | 6.9 | 2.0 | Appropriate | -1.97 | Appropriate |
| **5. Among patients treated with steroids alone in the hospital, it is appropriate to start ustekinumab after discharge if…** | | | | | | | | | | | | | | | |
| they have a low risk for AEs, the patient had a 𝘱𝘢𝘳𝘵𝘪𝘢𝘭 𝘳𝘦𝘴𝘱𝘰𝘯𝘴𝘦 with IV steroid rescue therapy, and is biologic/small molecule naïve prior to hospitalization. | 6.5 | 2.25 | 6.63 | 1.82 | Appropriate | -3.30 | Appropriate |  | 7 | 2.75 | 6.3 | 2.2 | Appropriate | 2.70 | Uncertain |
| they have a HIGH risk for AEs, the patient had a partial response with IV steroid rescue therapy, and is biologic/small molecule naïve prior to hospitalization. | 7 | 3 | 6.44 | 1.97 | Appropriate | -3.63 | Appropriate |  | 7 | 2.75 | 6.1 | 2.1 | Appropriate | 2.35 | Uncertain |
| they have a low risk for AEs, the patient had a partial response with IV steroid rescue therapy, and has been exposed to non-anti-IL12/23 biologic/small molecule therapy prior to hospitalization. | 7.5 | 2 | 6.88 | 1.93 | Appropriate | -3.08 | Appropriate |  | 7 | 1.75 | 6.9 | 2.0 | Appropriate | -0.71 | Appropriate |
| they have a HIGH risk for AEs, the patient had a partial response with IV steroid rescue therapy, and has been exposed to non-anti-IL12/23 biologic/small molecule therapy prior to hospitalization. | 7.5 | 2 | 7.00 | 1.59 | Appropriate | -3.08 | Appropriate |  | 7 | 1.75 | 6.8 | 2.0 | Appropriate | -0.71 | Appropriate |
| they have a low risk for AEs, the patient 𝘢𝘤𝘩𝘪𝘦𝘷𝘦𝘥 𝘳𝘦𝘮𝘪𝘴𝘴𝘪𝘰𝘯 with IV steroid rescue therapy, and is biologic/small molecule naïve prior to hospitalization. | 8 | 1 | 7.31 | 1.96 | Appropriate | -0.71 | Appropriate |  | 7.5 | 1 | 7.1 | 2.1 | Appropriate | -0.71 | Appropriate |
| they have a HIGH risk for AEs, the patient achieved remission with IV steroid rescue therapy, and is biologic/small molecule naïve prior to hospitalization. | 8 | 1.25 | 7.38 | 2.00 | Appropriate | -0.71 | Appropriate |  | 7.5 | 1.75 | 7.1 | 2.1 | Appropriate | -0.71 | Appropriate |
| they have a low risk for AEs, the patient achieved remission with IV steroid rescue therapy, and has been exposed to non-anti-IL12/23 biologic/small molecule therapy prior to hospitalization. | 8 | 1.25 | 7.94 | 0.77 | Appropriate | -0.02 | Appropriate |  | 8 | 1.75 | 7.9 | 0.9 | Appropriate | -0.71 | Appropriate |
| they have a HIGH risk for AEs, the patient achieved remission with IV steroid rescue therapy, and has been exposed to non-anti-IL12/23 biologic/small molecule therapy prior to hospitalization. | 8 | 0.5 | 7.88 | 1.02 | Appropriate | 0.00 | Appropriate |  | 8 | 1.75 | 7.8 | 1.1 | Appropriate | -0.44 | Appropriate |
| **6. Among patients treated with steroids alone in the hospital, it is appropriate to start ozanimod after discharge if…** | | | | | | | | | | | | | | | |
| they have a low risk for AEs, the patient had a 𝘱𝘢𝘳𝘵𝘪𝘢𝘭 𝘳𝘦𝘴𝘱𝘰𝘯𝘴𝘦 with IV steroid rescue therapy, and is biologic/small molecule naïve prior to hospitalization. | 5 | 4.25 | 5.25 | 2.49 | Uncertain | 1.61 | Uncertain |  | 4 | 3.75 | 4.6 | 2.4 | Uncertain | 0.76 | Uncertain |
| they have a HIGH risk for AEs, the patient had a partial response with IV steroid rescue therapy, and is biologic/small molecule naïve prior to hospitalization. | 5 | 3.25 | 4.75 | 2.44 | Uncertain | 0.97 | Uncertain |  | 4 | 2.75 | 3.9 | 2.0 | Uncertain | 0.52 | Uncertain |
| they have a low risk for AEs, the patient had a partial response with IV steroid rescue therapy, and has been exposed to non-S1P-receptor modulator biologic/small molecule therapy prior to hospitalization. | 5 | 3.25 | 5.00 | 2.16 | Uncertain | 0.57 | Uncertain |  | 5 | 1.75 | 4.4 | 1.8 | Uncertain | 0.32 | Uncertain |
| they have a HIGH risk for AEs, the patient had a partial response with IV steroid rescue therapy, and has been exposed to non-S1P-receptor modulator biologic/small molecule therapy prior to hospitalization. | 5 | 4 | 4.56 | 2.42 | Uncertain | 0.35 | Uncertain |  | 4.5 | 3 | 3.8 | 2.0 | Uncertain | 0.58 | Uncertain |
| they have a low risk for AEs, the patient 𝘢𝘤𝘩𝘪𝘦𝘷𝘦𝘥 𝘳𝘦𝘮𝘪𝘴𝘴𝘪𝘰𝘯 with IV steroid rescue therapy, and is biologic/small molecule naïve prior to hospitalization. | 7 | 3.5 | 5.88 | 2.45 | Appropriate | 2.35 | Uncertain |  | 6.5 | 3.25 | 6.1 | 2.3 | Appropriate | 10.00 | Uncertain |
| they have a HIGH risk for AEs, the patient achieved remission with IV steroid rescue therapy, and is biologic/small molecule naïve prior to hospitalization. | 5.5 | 3.25 | 5.13 | 2.53 | Uncertain | 1.73 | Uncertain |  | 5.5 | 4.5 | 4.9 | 2.6 | Uncertain | 1.35 | Uncertain |
| they have a low risk for AEs, the patient achieved remission with IV steroid rescue therapy, and has been exposed to non-S1P-receptor modulator biologic/small molecule therapy prior to hospitalization. | 6 | 2.25 | 6.25 | 1.57 | Uncertain | 2.35 | Uncertain |  | 6 | 2 | 5.9 | 1.7 | Uncertain | 1.36 | Uncertain |
| they have a HIGH risk for AEs, the patient achieved remission with IV steroid rescue therapy, and has been exposed to non-S1P-receptor modulator biologic/small molecule therapy prior to hospitalization. | 5.5 | 1.5 | 5.50 | 2.00 | Uncertain | 0.63 | Uncertain |  | 5.5 | 1.75 | 5.1 | 2.0 | Uncertain | 0.63 | Uncertain |
| **7. Among patients treated with steroids alone in the hospital, who were exposed to anti-TNF therapy prior to the hospitalization, it is appropriate to start tofacitinib after discharge if…** | | | | | | | | | | | | | | | |
| they have a low risk for AEs, and the patient had a 𝘱𝘢𝘳𝘵𝘪𝘢𝘭 𝘳𝘦𝘴𝘱𝘰𝘯𝘴𝘦 with IV steroid rescue therapy. | 7.5 | 1.25 | 7.25 | 1.81 | Appropriate | -0.71 | Appropriate |  | 7 | 1.75 | 7.2 | 2.0 | Appropriate | -0.71 | Appropriate |
| they have a HIGH risk for AEs, and the patient had a partial response with IV steroid rescue therapy. | 7 | 2.25 | 6.00 | 1.97 | Appropriate | 2.35 | Uncertain |  | 6 | 2 | 5.5 | 1.8 | Uncertain | 1.36 | Uncertain |
| they have a low risk for AEs, and the patient 𝘢𝘤𝘩𝘪𝘦𝘷𝘦𝘥 𝘳𝘦𝘮𝘪𝘴𝘴𝘪𝘰𝘯 with IV steroid rescue therapy. | 8 | 2 | 7.75 | 1.18 | Appropriate | -0.71 | Appropriate |  | 7 | 1.75 | 7.6 | 1.0 | Appropriate | -0.71 | Appropriate |
| they have a HIGH risk for AEs, and the patient achieved remission with IV steroid rescue therapy. | 6 | 2 | 6.00 | 1.86 | Uncertain | 7.64 | Uncertain |  | 6 | 1.75 | 5.6 | 1.8 | Uncertain | 0.51 | Uncertain |
